# Estimation of COVID-19 cases prevented by vaccination in California

**DOI:** 10.1101/2022.02.08.22269319

**Authors:** Sophia Tan, Hailey Park, Isabel Rodríguez-Barraquer, George W. Rutherford, Kirsten Bibbins-Domingo, Nathan C. Lo

## Abstract

**Importance:** Despite widespread vaccination against COVID-19 in the United States, there are limited empirical data quantifying the public health impact in the population.

**Objective:** To estimate the number of cases of COVID-19 averted due to COVID-19 vaccination

**Design, Setting, and Participants:** The California Department of Public Health (CDPH) provided person-level data on COVID-19 cases and COVID-19 vaccine administration. To estimate the number of COVID-19 cases that would have occurred in the vaccine era in absence of vaccination, we applied a statistical model that estimated the relationship of COVID-19 cases in the pre-vaccine era between the unvaccinated age group (<12 years) and vaccine-eligible groups (≥12 years) to COVID-19 case data after the start of vaccination. The primary study outcome was the difference between predicted number of COVID-19 cases in absence of vaccination and observed COVID-19 cases with vaccination. As a sensitivity analysis, we developed a second independent model that estimated the number of vaccine-averted COVID-19 cases by applying published data on vaccine effectiveness to data on COVID-19 vaccine administration and estimated risk of COVID-19 over time.

**Intervention:** COVID-19 vaccination

**Main Outcomes and Measures:** COVID-19 cases

**Results:** There were 4,585,248 confirmed COVID-19 cases in California from January 1, 2020 to October 16, 2021, during which 27,164,680 vaccine-eligible individuals ≥12 years were reported to have received at least 1 dose of a COVID-19 vaccine in the vaccine era (79.5% of the eligible population). We estimated that 1,523,500 [95% prediction interval (976,800-2,230,800)] COVID-19 cases were averted and there was a 34% [95% prediction interval (25-43)] reduction in cases due to vaccination in the primary model. Approximately 66% of total cases averted occurred after the delta variant became the dominant strain of SARS-CoV-2 circulating in California. Our alternative model identified comparable findings.

**Conclusions and Relevance:** This study provides robust evidence on the public health impact of COVID-19 vaccination in the United States and further supports the urgency for continued vaccination.

**Key Points:** *Question:* How many COVID-19 cases have been prevented by COVID-19 vaccination in California?

*Findings:* In this empirical analysis of California using data from the Department of Public Health, we estimated that COVID-19 vaccination has prevented over 1.5 million COVID-19 cases from the introduction of vaccination through October 16, 2021.

*Meaning:* These findings support that COVID-19 vaccination had a large public health impact in California in terms of averted cases of COVID-19 and can be generalized across the United States.

## Introduction

COVID-19 has caused substantial morbidity, mortality, and socioeconomic disruption and disparity in the United States and globally. The COVID-19 vaccine has been a key tool for public health control of COVID-19, alongside public health measures for universal masking and social distancing^1,2^. In the United States, implementation of COVID-19 vaccination took place in a phased approach, guided by recommendations from the US Centers for Disease Control and Prevention Advisory Committee on Immunization Practices (ACIP). The ACIP recommendations prioritized vaccination based on risk of infection. Phase 1a, 1b, and 1c of vaccination included healthcare personnel, essential workers, adults ≥65 years, and individuals with high-risk conditions. Phase 2 included vaccination of the general population ≥16 years^3^. Initiation of Phase 1a vaccination against COVID-19 began in December 2020^3,4^, signaling the initiation of widespread vaccination in the United States.

Three vaccines are authorized in the United States: 1) BNT162b2-mRNA from Pfizer/BioNTech (Pfizer/BioNTech); 2) mRNA-1273 from Moderna (Moderna); and 3) single dose Ad26.COV2.S from Janssen (Janssen). The pivotal trials on these vaccines demonstrated high efficacy against clinical disease, hospitalization, and death; efficacy against symptomatic infection was 95%, 94%, and 66% in the Pfizer/BioNTech, Moderna, and Janssen vaccines respectively^5–7^. Furthermore, published data on the real-world effectiveness have demonstrated similar protection against clinical disease, with some waning for the Pfizer/BioNTech BNT162b2-mRNA and Janssen vaccine^8^ and some differential vaccine effectiveness by infecting variant^9–13^. However, despite widespread implementation of COVID-19 vaccinations in the United States, there are limited data to estimate the overall public health impact of vaccination on averted cases of COVID-19.

This article reports on the public health impact of COVID-19 vaccination by estimating the number of COVID-19 cases averted in the first 10 months of vaccination, using the representative case example of California given the large population, geographic size, and epidemiologic variation within the state.

## Methods

We developed two independent statistical modeling approaches to estimate the number of averted COVID-19 cases due to direct effects of vaccination. In this analysis, we used person-level COVID-19 case data from the California Department of Public Health (CDPH) and public data on COVID-19 vaccination.

### Data

We obtained de-identified person-level case data for confirmed COVID-19 cases in California from January 1, 2020, to October 16, 2021, from CDPH. A case of COVID-19 was defined as a person whose positive SARS-CoV-2 molecular test was reported to the state, including both symptomatic cases and asymptomatic infections^14^. We excluded data on persons with missing age data (<1%).

We obtained publicly available CDPH vaccine administration data in four age groups (12-17 years, 18-49 years, 50-64 year, ≥65 years) from July 27, 2020 to October 16, 2021^15^. These age groupings were based upon vaccine eligibility^16^. We excluded vaccination in children 5-11 years old (vaccine coverage <0.01%) since they were ineligible over the study period^17,18^. We additionally excluded vaccination that occurred before start of Phase 1a of vaccination (<0.01%) or had missing age information (<0.02%). We aggregated both COVID-19 case and vaccination data from daily counts to weekly counts beginning January 1, 2020. This project was approved by the institutional review board at the University of California, San Francisco.

### Study outcomes

The primary study outcome was COVID-19 cases averted due to the direct effects of COVID-19 vaccination. We also estimated the relative reduction in COVID-19 cases due to vaccination (see Appendix). Secondary study outcomes include averted COVID-19 cases and relative reduction of cases by age group. Study outcomes were chosen based on public health relevance. Sufficient data were not available to estimate vaccination impact on COVID-19 related deaths or hospitalizations.

### Statistical analysis

We used two independent modeling approaches to estimate the number of averted COVID-19 cases due to vaccination in the vaccine era from November 29, 2020 to October 16, 2021. We defined the start of the vaccine era based on the approximate start of Phase 1a of COVID-19 vaccination in California^4^. We chose to develop multiple estimation procedures that relied on different assumptions to improve the reliability and robustness of our study findings. Analytic code is available on GitHub^19^. All analyses were conducted in R (version 3.6.0).

#### Primary statistical model

In the primary model, we predicted the number of COVID-19 cases that would have otherwise occurred over time if vaccines were never available. This modeling approach used the unvaccinated population to serve as a proxy for overall force of infection over time in the vaccine-eligible population. We defined our unvaccinated population as all children <12 years as they were ineligible for vaccination over the entire study period.

We used quasi-Poisson regression models to estimate the association between the log-transformed number of weekly COVID-19 cases in the unvaccinated population (<12 years) and the number of weekly cases in each of the four vaccine-eligible populations (12-17 years, 18-49 years, 50-64 year, ≥65 years) during the pre-vaccine era at the state level. The pre-vaccine era was defined as May 31, 2020 to November 28, 2020 to provide 6 months of data for model calibration. We fit separate models for each of the four age-groups of the vaccine-eligible population.

We then applied each calibrated model to the observed COVID-19 cases in the unvaccinated population (<12 years) in the vaccine era (November 29, 2020 to October 16, 2021) to predict the number of COVID-19 cases that would have occurred in each age-group in the absence of vaccination. We computed the weekly difference between the predicted number of COVID-19 cases in absence of vaccination with the observed number of COVID-19 cases from the CDPH dataset (see Appendix). We summed the differences across age groups to estimate the total number of averted COVID-19 cases due to the direct protection of COVID-19 vaccination in California. We reported 95% prediction intervals (95% PI) for study estimates. This model relied on the assumption that the relative risk of COVID-19 diagnosis remained constant over time between unvaccinated population (<12 years) and vaccine-eligible age groups. This model did not explicitly account for person-level vaccination.

#### Alternative statistical model

In the second model, we used published data on vaccine effectiveness and estimated risk of COVID-19 in the vaccine era (November 29, 2020 to October 16, 2021) to predict the number of COVID-19 cases averted due to the direct protection of vaccination. First, we estimated state-level weekly incidence of COVID-19 cases (defined as total cases per 100,000 susceptible persons) beginning January 1, 2020 (earliest available data for COVID-19 in California) in each of the four vaccine-eligible populations (12-17 years, 18-49 years, 50-64 year, ≥65 years). We estimated the fraction of the population susceptible to infection over time by age group based on natural infection or vaccination. For natural immunity, we used our data on reported COVID-19 cases and applied published estimates of age-specific clinical fractions to estimate the total number of infections (including sub-clinical infections) (see Appendix)^20^. We made the simplifying assumption that natural infection provided perfect immunity. For vaccine-induced immunity, we used data on vaccine administration and published data on vaccine effectiveness^5–8^. We estimated vaccine-induced immunity and waning specific to each of the COVID-19 vaccines (Pfizer/BioNTech, Moderna, Janssen) over time (see Appendix).

We applied the age-specific estimated risk of COVID-19 to the fraction of the corresponding age group susceptible to infection, removing vaccine-induced protection and thus simulating a scenario without vaccination. This estimation was conducted in the vaccine era (November 29, 2020 – October 16, 2021). We estimated 95% uncertainty intervals (95% UI), which incorporated uncertainty in estimates of vaccine effectiveness over time and the age-specific asymptomatic fraction generated through Monte Carlo simulation (see Appendix). This approach did not assume a fixed relationship in COVID-19 cases between the unvaccinated and vaccine-eligible groups. This model did assume complete reporting of both case and vaccination data and a constant asymptomatic fraction of infection over time.

### Sensitivity Analyses

We conducted several sensitivity analyses to evaluate the effects of varying assumptions and parameters in the model. We estimated the relative reduction in COVID-19 cases due to vaccination, while accounting for differences in timing of age-specific vaccine eligibility in both models (see Appendix). This is in contrast to our base case assumption where vaccination is measured from the start of Phase 1A of vaccination, which provides an underestimate of the impact of vaccination since most of the general population was not yet eligible for vaccine. In the primary model, we varied the calibration period and the impact of using other age group definitions for the unvaccinated population. In the alternative model, we varied vaccine effectiveness estimates to simulate the study finding under reduced vaccine effectiveness to the delta variant (see Appendix).

## Results

### Descriptive data

There were 4,588,146 reported COVID-19 cases in California from January 1, 2020, to October 16, 2021. We included 4,585,248 COVID-19 cases and excluded 2,898 (0.06%) due to missing age data. 3,276,260 COVID-19 cases (71%) occurred after November 28, 2020 (Phase 1a of COVID-19 vaccination in California). There were 899,510 confirmed cases of COVID-19 after the delta variant became a prominent circulating strain of SARS-CoV-2 in June 2021 (Figure 1)^21^.

**Figure 1:**
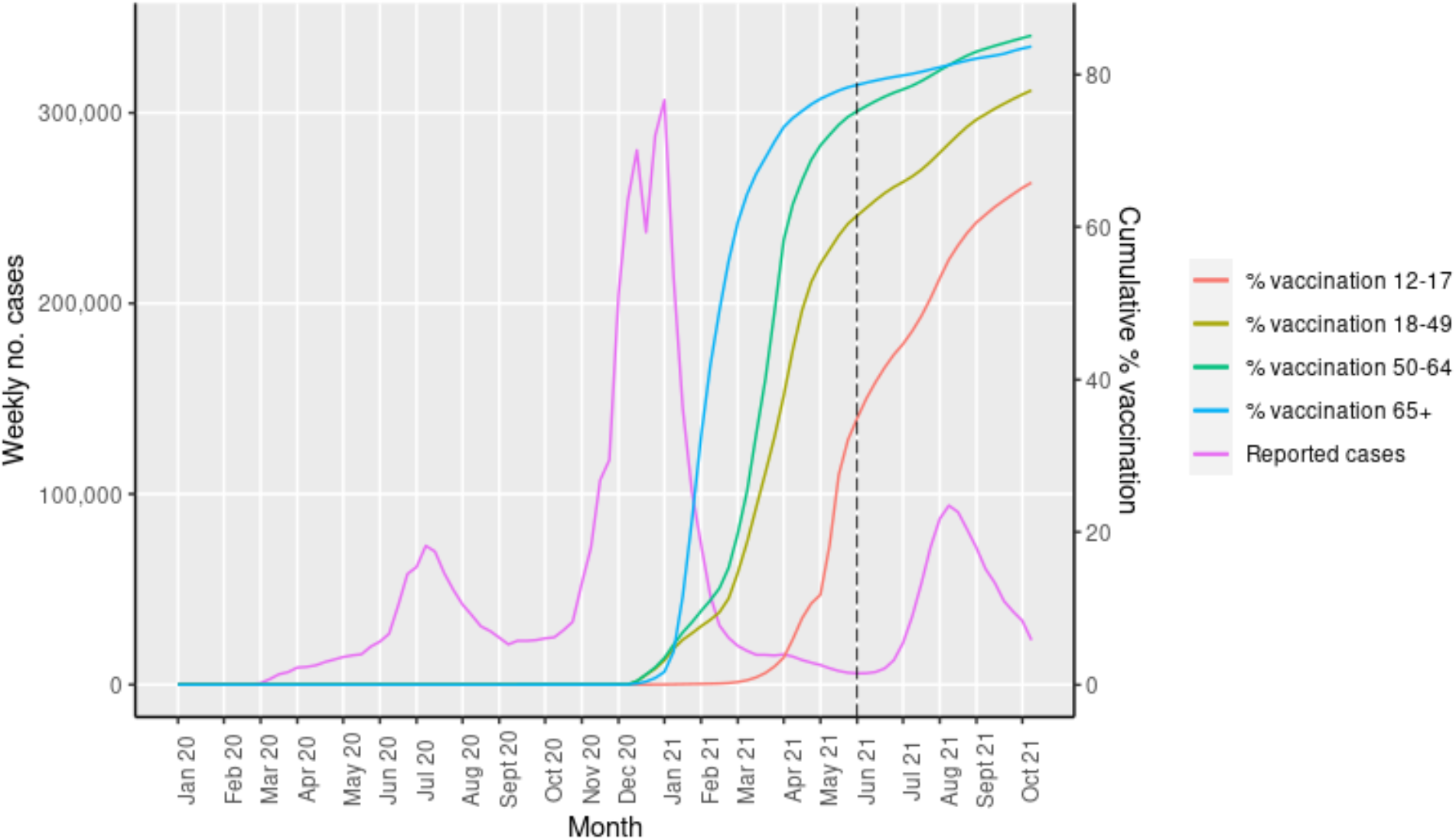
COVID-19 cases and vaccination over time in California. Data on COVID-19 cases were obtained from CPDH for the period of January 1, 2020 to October 16, 2021. Weekly absolute COVID-19 cases were plotted (purple). We plotted cumulative coverage of COVID-19 vaccination by age-group using publicly available data from January 1, 2020 to October 16, 2021. We defined date of vaccination as the date of first vaccine dose receipt in persons who received at least 1 dose of a COVID-19 vaccine (Pfizer/BioNTech, Moderna, Janssen). The dashed line (black) represents introduction of the delta variant in California.

Between November 29, 2020 and October 16, 2021, 27,164,680 (79.5%) persons ≥12 years were reported to have received at least 1 dose of a COVID-19 vaccine in California. We excluded 2,022 individuals (<0.01%) that received vaccines before November 29, 2020 (Figure 1). Approximately 57% of vaccine-eligible individuals received the Pfizer/BioNTech vaccine, 36% received the Moderna vaccine, and 7% received the Janssen vaccine. We excluded 3,924 (<0.02%) individuals with missing age from the vaccination data.

### Primary model results

We observed good model fit over the calibration period for the primary model in the pre-vaccine era (May 31, 2020 to November 28, 2020) (Figure 2). We estimated that 1,523,500 [95% PI (976,800-2,230,800)] COVID-19 cases were averted due to COVID-19 vaccination (Table 1 and Figure 3), which corresponded with a 34% [95% PI (25-43)] overall reduction in cases in the vaccine-eligible population after the start of Phase 1a of vaccination (Table 1) and a 46% [95% PI (43-49)] reduction in cases when taking into account age-specific differences in vaccine eligibility (eTable 1). The 12-17 years, 18-49 years, 50-64 years, and ≥65 years populations experienced 15%, 36%, 34% and 30% reductions in COVID-19 cases respectively after the start of Phase 1a of vaccination (Table 1 and eFigure 1) and experienced greater reductions when accounting for age-specific eligibility over time (eTable 1). Approximately 1,005,500 [95% PI (809,300-1,230,450)] COVID-19 cases (66% of total cases averted) were averted after the delta variant became the dominant strain of SARS-CoV-2 circulating in California in June 2021^21^.

**Figure 2:**
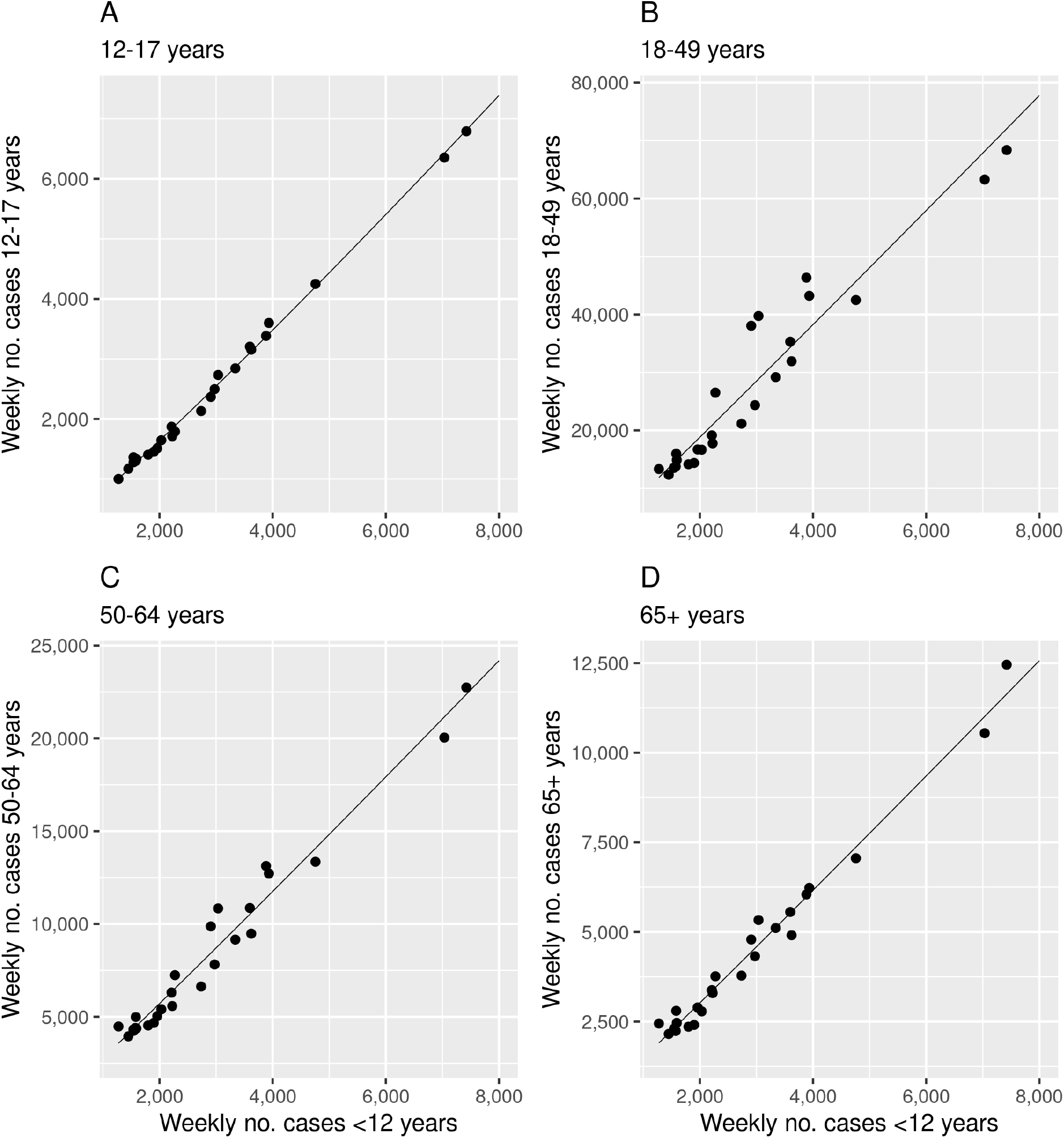
Model calibration of primary model for relationship between COVID-19 cases in the unvaccinated and vaccine-eligible age groups in the pre-vaccine era. In our primary model, we plotted weekly cases in the unvaccinated population (<12 years) and each vaccine-eligible age group (12-17 years in panel A, 18-49 years in panel B, 50-64 years in panel C, and ≥65 years in panel D) before Phase 1a of vaccination. We fit quasi-Poisson models between log-transformed weekly cases in the unvaccinated population and weekly cases in each vaccine-eligible age group (black line) and observed good model fit in each age group. The strong linear relationship suggests a good model fit during the calibration. We used the calibrated models to make predictions on COVID-19 cases in the vaccine era under the scenario of no vaccination.

**Table 1:** Comparison of primary and alternative model to estimate the number of COVID-19 cases averted due to vaccination in California by age group.

|  | Age group (years) | Observed cases | Predicted COVID-19 cases (95% PI or UI) | Averted COVID-19 cases (95% PI or UI) | Relative reduction in % (95% PI or UI) |
| --- | --- | --- | --- | --- | --- |
| **Primary model** | Total ($\geq 12$) | 2,983,152 | 4,506,630 (3,959,910, 5,071,680) | 1,523,500 (976,800, 2,230,800) | 34 (25, 43) |
|  | 12-17 | 238,031 | 278,960 (268,300, 290,120) | 40,930 (30,270, 52,090) | 15 (11, 18) |
|  | 18-49 | 1,829,625 | 2,866,350 (2,444,720, 3,412,980) | 1,036,700 (615,100, 1,588,300) | 36 (25, 46) |
|  | 50-64 | 588,553 | 894,900 (810,040, 1,003,720) | 306,300 (221,500, 415,200) | 34 (27, 41) |
| | $\geq 65$ | 326,943 | 466,410 (436,850, 507,170) | 139,500 (109,900, 180,200) | 30 (25, 36) |
| **Alternative model** | Total ($\geq 12$) | 2,983,152 | 4,385,300 (4,175,200, 4,598,700) | 1,402,100 (1,192,100, 1,615,600) | 32 (29, 35) |
|  | 12-17 | 238,031 | 316,790 (304,170, 328,640) | 78,760 (66,140, 90,610) | 25 (22, 28) |
|  | 18-49 | 1,829,625 | 2,640,320 (2,527,610, 2,751,860) | 810,700 (697,990, 922,240) | 31 (28, 34) |
|  | 50-64 | 588,553 | 909,830 (857,940, 963,950) | 321,280 (269,390, 375,390) | 35 (31, 39) |
| | $\geq 65$ | 326,943 | 518,340 (485,510, 554,280) | 191,390 (158,570, 227,340) | 37 (33, 41) |

**Figure 3:**
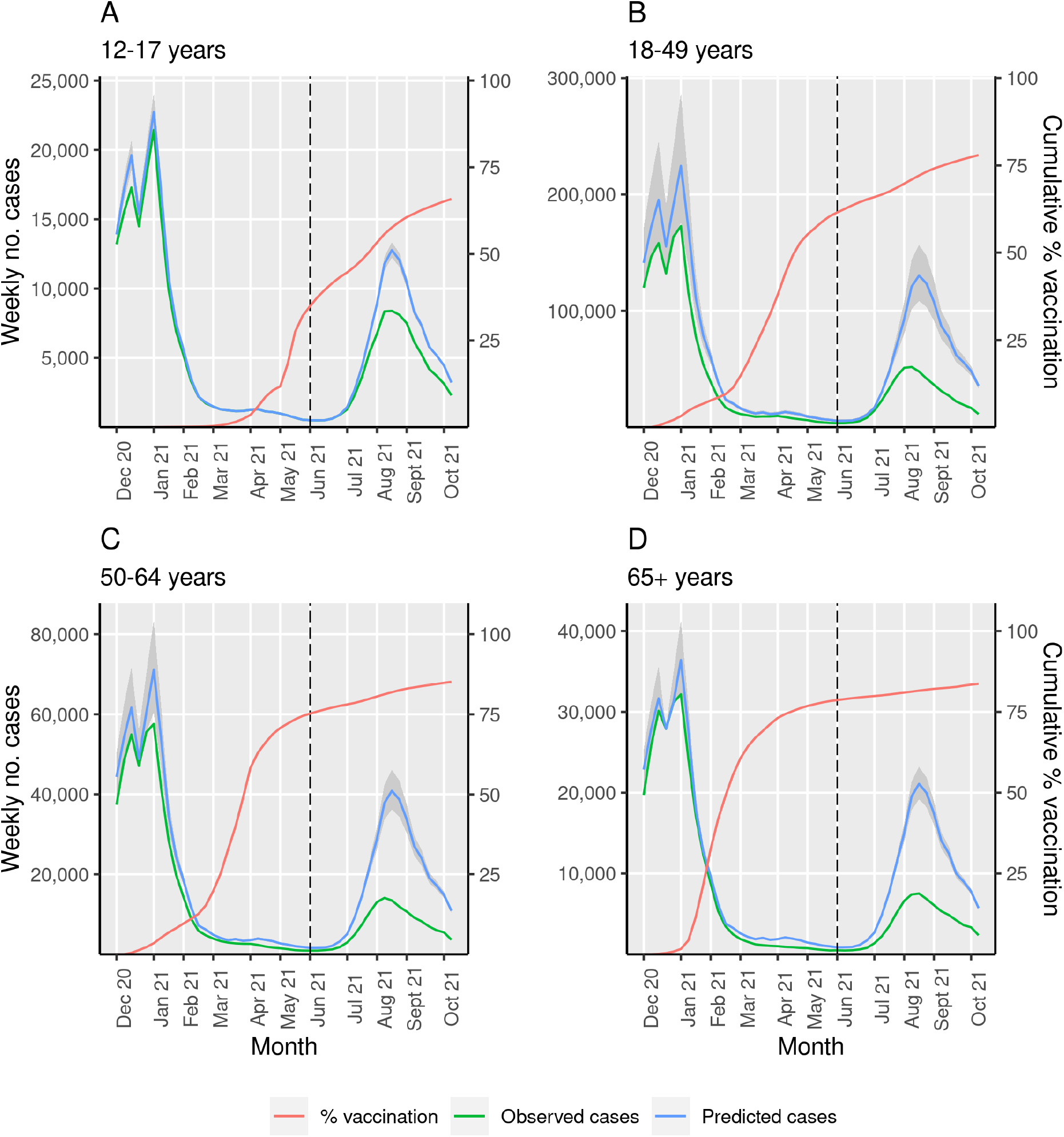
Primary model estimating averted COVID-19 cases due to COVID-19 vaccination in California by age group. We plotted observed COVID-19 cases (green) and vaccine coverage of at least one dose of a COVID-19 vaccine (red) over time in four age groups that represent the vaccine-eligible population: 12-17 years (A), 18-49 years (B), 50-64 years (C) and ≥65 years (D). In the primary analysis, we estimated the association between cases in the unvaccinated group (<12 years) and each vaccine-eligible age-group before Phase 1a of vaccination to predict COVID-19 cases in each vaccine-eligible age-group in absence of vaccination (blue) with prediction intervals in grey. The dashed line (black) represents introduction of the delta variant in California. The difference between predicted COVID-19 cases without vaccination (blue) and observed COVID-19 cases with vaccine (green) represents the averted COVID-19 cases due to vaccination.

We performed sensitivity analyses on the primary model, including testing alternative age definitions for the vaccine-eligible population and date to define vaccine introduction, with overall similar findings (see Appendix).

### Alternative model results

In our alternative modeling approach, we estimated that 1,402,100 [95% UI (1,192,100-1,615,600)] were averted due to vaccination from November 29, 2020 to October 16, 2021 (Table 1 and Figure 4). We estimated that vaccination contributed to a 32% [95% UI (29-35)] reduction in cases in the population ≥12 years after the start of vaccination (Table 1) and a 50% [95% UI (47-53)] reduction in cases when accounting for age-specific differences in vaccine-eligibility (eTable1). The populations 12-17 years, 18-49 years, 50-64 years, and ≥65 years experienced 25%, 31%, 35% and 37% reductions in COVID-19 cases after the start of Phase 1a of vaccination, respectively (Table 1 and eFigure 2) and greater reductions in cases when accounting for differences in vaccine-eligibility (eTable 1). We estimated 90% of averted cases were prevented after the introduction of the delta variant.

**Figure 4:**
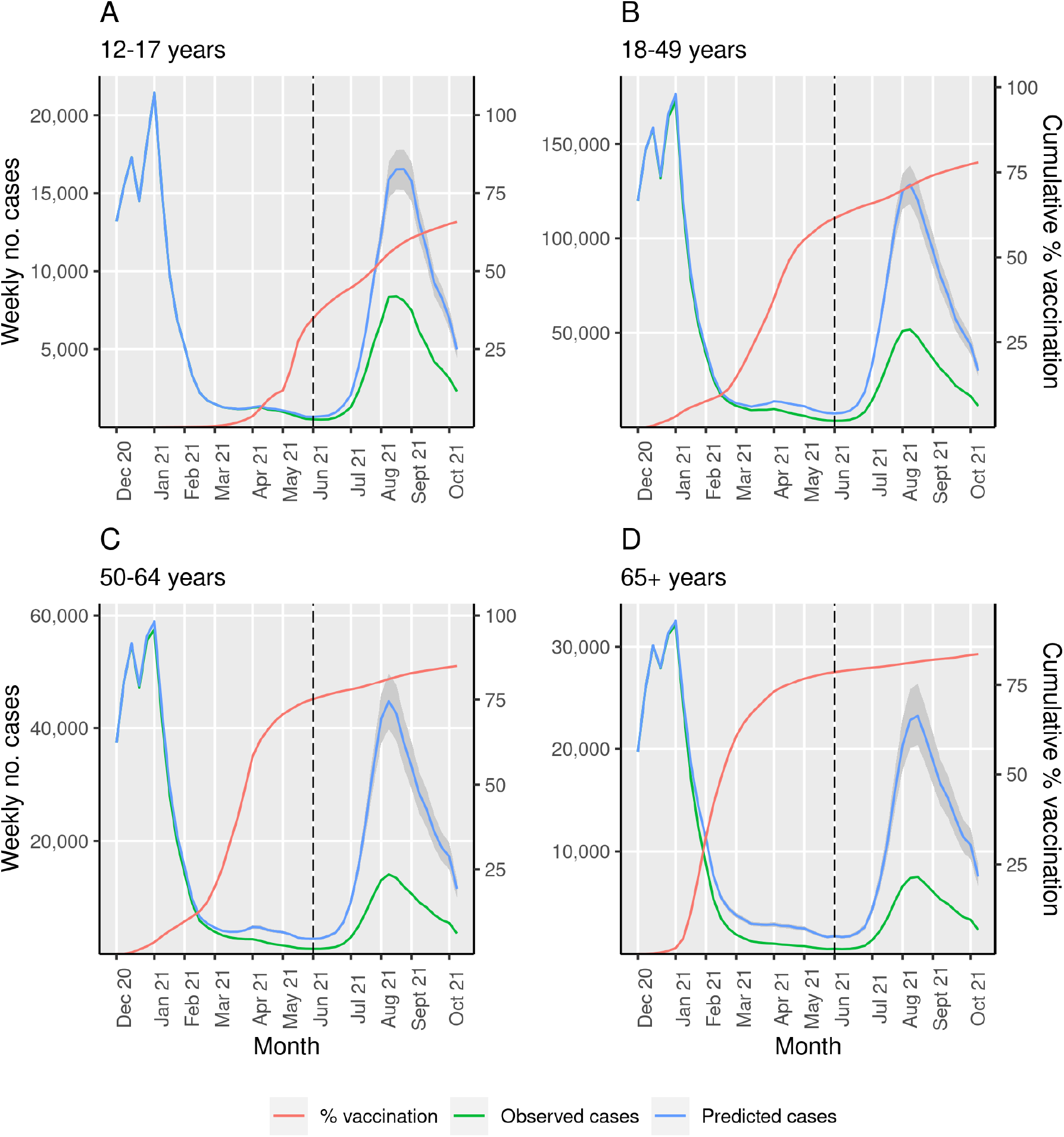
Alternative model estimating averted COVID-19 cases due to COVID-19 vaccination in California by age group. This analysis used an alternative modeling approach. We estimated the COVID-19 incidence in each vaccine-eligible age group over time, accounting for natural and vaccine-induced immunity. We used this age-specific COVID-19 incidence to predict weekly cases in the vaccine-eligible group, under the scenario of no vaccination (blue) with uncertainty intervals in grey. We plotted observed COVID-19 cases (green) as well as vaccine coverage of at least one dose of a COVID-19 vaccine (red) over time in four separate age groups that represent the vaccine-eligible population: 12-17 years (A), 18-49 years (B), 50-64 years (C) and ≥65 years (D). The difference between predicted COVID-19 cases without vaccination (blue) and observed COVID-19 cases with vaccine (green) represents the averted COVID-19 cases due to vaccination. The dashed line (black) represents introduction of the delta variant in California.

In sensitivity analysis, we obtained comparable results when accounting for the possibility of reduced vaccine effectiveness against the delta variant; we estimated that 1,106,300 [95% UI (913,300-1,308,650)] cases were prevented, and there was a 27% reduction [95% UI (23, 30)] in cases (eTable 4). In this sensitivity analysis, 87% of averted COVID-19 cases were prevented during widespread transmission of the delta variant.

## Discussion

In this study, we found that over 1.5 million COVID-19 cases were prevented due to protection by COVID-19 vaccination programs over the first 10 months of widespread vaccination in California. Vaccination contributed to an over 30% reduction in cases. These study findings are strengthened by our second modeling approach which had comparable findings. This study reports predominately on the direct effects of vaccination and is not able to capture the indirect effects of vaccination (i.e., averted cases due to reduced SARS-CoV-2 transmission), meaning the overall impact of vaccination is likely larger than estimated. These findings support that COVID-19 vaccination had a large public health impact in California in terms of averted cases of COVID-19, which likely generalizes across the United States.

A large number of COVID-19 cases were prevented after the introduction of the SARS-CoV-2 delta variant in California, which is more infectious than other previously identified variants of SARS-CoV-2^22^. The primary model and alternative model estimated over 66% and 90% of averted cases occurred after the delta variant became the prominent variant of SARS-CoV-2 circulating in California respectively. This suggests the burden of COVID-19 cases and more severe outcomes, such as hospitalizations and deaths, would likely have been significantly higher in the absence of vaccination. We found a similar impact of vaccination when additionally considering reduced effectiveness of vaccines against the delta variant.

We estimated over a 30% reduction in COVID-19 cases in the vaccine-eligible population in California from the start of Phase 1a of vaccination. This provided a conservative estimate of the true reduction since vaccine eligibility across age groups varied over time. When accounting for differences in vaccine eligibility by age, we found over a 45% reduction in COVID-19 cases. However, this still likely represents an underestimate as early vaccine eligibility was determined by occupational and health risk, which our data was unable to fully capture.

A key strength of the analysis is the development of two distinct modeling approaches to improve reliability of the study findings. While each analysis relied on limiting assumptions, the assumptions were non-overlapping and overall findings were similar. The primary model estimated 1.5 million averted COVID-19 cases and a 34% reduction in cases due to vaccination, and the alternative model found over 1.4 million averted cases and a 32% reduction in cases (Table 1). Notably, the primary modeling approach predicted a greater number of cases averted and greater reduction in cases early in the vaccine era (Figures 4 and eFigure 1). The alternative modeling approach predicted greater vaccine impact after the widespread circulation of the delta variant (Figures 5 and eFigure 2). The alternative modeling approach predicted more cases averted and greater reduction in cases in the populations 12-17 years and ≥65 years than the primary modeling approach (Table 1).

Our primary modeling approach relied on the key assumption that COVID-19 cases in the unvaccinated (<12 years) population remained a robust predictor of cases in each vaccine-eligible group over time. We assumed that the relative risk for SARS-CoV-2 infection between the vaccine ineligible (<12 years) and vaccine-eligible populations was stable over time, as well as stable testing practices within and between these age groups. Relative risk and testing practices likely changed over time and differentially between groups, especially as children returned to schools and had access to increased SARS-CoV-2 testing in the fall of 2021^23^. To address this, we developed the alternative modeling approach that did not make this assumption and ultimately had similar study findings. Power limitations prevented our analysis from studying other clinical outcomes such as hospitalizations or deaths.

Our alternative modeling approach used the susceptibility profile of the vaccine-eligible population over time to estimate the COVID-19 cases prevented by vaccination. The key assumption in the alternative model was complete reporting of COVID-19 cases and vaccination, although this assumption was relaxed for estimating the relative reduction outcome. Similarly, we assumed the symptomatic fraction of reported SARS-CoV-2 infections remained constant over time, even with the introduction of vaccines (see Appendix). An increase in asymptomatic screening over time would bias the study to overestimate the public health impact of vaccination, while an increase in the asymptomatic fraction of infection over time due to vaccine effectiveness would bias the study to underestimate the public health impact of vaccination.

In conclusion, we found that COVID-19 vaccination had a substantial public health impact by reducing COVID-19 cases in California. This study provides evidence on the public health impact of COVID-19 vaccination in the United States, and further supports the urgency for continued vaccination.

## Data Availability

Data on COVID-19 vaccination is publicly available online, and data on COVID-19 cases is available on request to CDPH. Analytic code is available online.

https://github.com/sophttan/ca-vaccine-impact

https://data.chhs.ca.gov/dataset/vaccine-progress-dashboard/resource/faee36da-bd8c-40f7-96d4-d8f283a12b0a

## Acknowledgments

We thank the California Department of Public Health for sharing the data used in this article and appreciate all the individuals involved in data collection and curation. We specifically appreciate assistance from the CDPH COVID-19 Data Processing and Informatics Section and COVID-19 Modeling Team. We would also like to thank all those involved in the ongoing response to the COVID-19 pandemic in California.

## Authorship contribution

Ms. Sophia Tan and Dr Nathan Lo had full access to all the data in the study and take responsibility for the integrity of the data and the accuracy of the data analysis.

Study concept and design: ST, IRB, RS, NCL

Statistical analysis: ST, NCL

Acquisition, analysis, or interpretation of data: HP, ST, IRB, RS, NCL

First draft of the manuscript: ST, NCL

Critical revision of the manuscript: All authors

Contributed intellectual material and approved final draft: All authors

## Declaration of interests and financial disclosures

None

## Funding/Support

California Department of Public Health

## Role of the Funding Organization or Sponsor

The funding organizations had no role in the design and conduct of the study; collection, management, analysis, and interpretation of the data; and preparation, review, or approval of the manuscript; and decision to submit the manuscript for publication. This work represents the
viewpoints of the authors alone and not necessarily those of the California Department of Public Health.

## Previous presentations

None.

## Notes

### Competing Interest Statement

The authors have declared no competing interest.

### Funding Statement

This study was funded by the California Department of Public Health. The funding organizations had no role in the design and conduct of the study; collection, management, analysis, and interpretation of the data; and preparation, review, or approval of the manuscript; and decision to submit the manuscript for publication. This work represents the viewpoints of the authors alone and not necessarily those of the California Department of Public Health.

### Author Declarations

This study was approved by the institutional review board at the University of California, San Francisco.

