## Supplemental Materials for "Estimation of COVID-19 cases prevented by vaccination in California"

### Technical Appendix

In this appendix, we provide further methodologic detail on the model structure and statistical analysis.

<u>Study outcomes</u>

*Relative reduction of cases*

We estimated the relative reduction of cases in the entire vaccine-eligible population (≥12 years) and each age group (12-17 years, 18-49 years, 50-64 years, and ≥65 years) after the start of Phase 1A of vaccination (November 29, 2020) and, alternatively, when accounting for age-specific eligibility over time (see Sensitivity analyses). The formula for percentage reduction in COVID-19 cases over a fixed period of time is as follows:

$$\%\, reduction\ in\ clinical\ cases = \frac{predicted\ cases - observed\ cases}{predicted\ cases} \times 100$$

<u>Primary model: Additional methodology</u>

We defined the lower bound for the number of weekly averted COVID-19 cases as zero based on bioplausibility.

<u>Alternative model: Additional methodology</u>

*Model of natural immunity*

We assumed that natural infection provided perfect immunity without waning. We assumed complete reporting of COVID-19 cases. We estimated total infections in the unvaccinated and each vaccine-eligible age group (<12 years, 12-17 years, 18-49 years, 50-64 years, ≥65 years) using literature estimates of the asymptomatic proportion by age (<19 years, 19-59 years, ≥60 years)[1]. We used the reported means and 95% confidence intervals of each age-specific asymptomatic fraction of infection to fit optimal beta distributions. The mean and the fitted shape parameters of each distribution are shown in Table A1.

**Table A1: Mean and fitted parameters for the distribution of asymptomatic fraction of SARS-CoV-2 infection by age group[1]**

| Age group (years) | | Asymptomatic proportion | |
|---|---|---|---|
| Primary age groups | Age subgroups | Mean | Beta distribution (α, β) |
| <12 | <12 | 0.47 | Beta (19.3, 21.8) |
| 12-17 | 12-17 | 0.47 | Beta (19.3, 21.8) |
| 18-49 | 18 | 0.47 | Beta (19.3, 21.8) |
| | 19-49 | 0.32 | Beta (22.4, 46.7) |
| 50-64 | 50-59 | 0.32 | Beta (22.4, 46.7) |
| | 60-64 | 0.2 | Beta (17.6, 69.2) |
| ≥65 | ≥65 | 0.2 | Beta (17.6, 69.2) |

For each age group $a$, we estimated the total infections at week $t$ using the following formula:

$$I_{a,t} = \sum_{i \epsilon a} \frac{C_{i,t}}{1 - p_i}$$

$I_{a,t} = \ total\ infections$
$C_{i,t} = observed\ COVID - 19\ cases$
$p_i = asymptomatic\ proportion$

*Model of vaccine-induced immunity*
We modeled vaccine effectiveness (against clinical disease) and waning immunity on a person-level based on vaccine (Pfizer/BioNTech, Moderna, Janssen) and the number of doses received. We assumed six possible vaccination scenarios: 1) Pfizer/BioNTech single dose; 2) Pfizer/BioNTech two doses; 3) Moderna single dose; 4) Moderna two doses; 5) Janssen single dose; and 6) unvaccinated.

We used published literature to estimate the vaccine effectiveness in each scenario over time, assuming instantaneous onset of protection and waning immunity at various time points[2–6]. We fit beta distributions using the published mean and 95% confidence intervals of each estimate of vaccine effectiveness (see Table A2). We made the simplifying assumption that all individuals who received two doses of the Pfizer/BioNTech vaccine received their second dose three weeks after their first dose and all individuals that received two doses of the Moderna vaccine received their second dose four weeks after receiving their first dose based on published literature[7]. We did not account for potential differences in vaccine effectiveness by age, which is broadly supported by literature[6,8,9]. We accounted for possible changes in vaccine effectiveness against the highly infectious delta variant of SARS-CoV-2 as a sensitivity analysis (see Sensitivity analyses) but did not account for variant specific effectiveness in the main analysis. Average vaccine effectiveness and waning over time is shown in Figure A1, and the distributions of vaccine effectiveness are shown in Table A2.

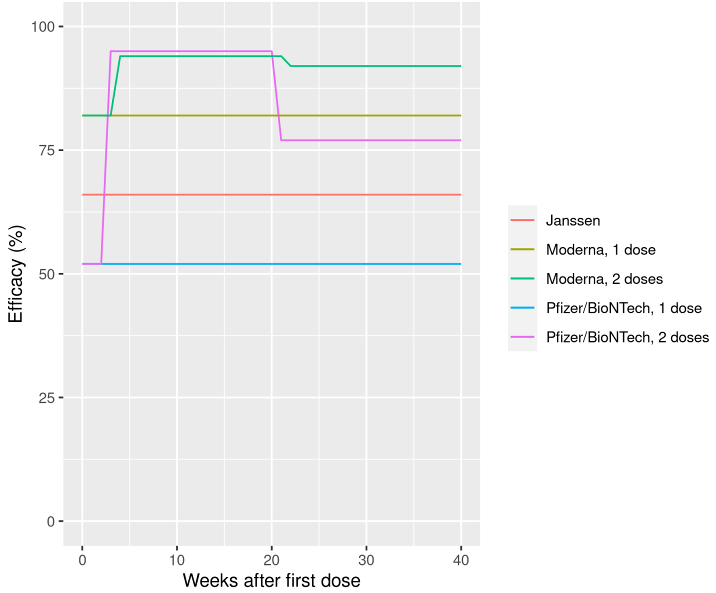

**Figure A1: Mean COVID-19 vaccine effectiveness against clinical infection over time by manufacturer and number of doses**

**Table A2: COVID-19 vaccine effectiveness by manufacturer with mean and distribution**

| Vaccine | Vaccine period | Vaccine effectiveness | |
|---|---|---|---|
| | | Mean | Beta distribution (α, β) |
| Pfizer/BioNTech | Between 1st and 2nd dose | 0.52[3] | Beta (12, 11.9) |
| | ≤17 weeks after 2nd dose | 0.95[3] | Beta (128.5, 7.2) |
| | >17 weeks after 2nd dose | 0.77[5] | Beta (71.3, 22.1) |
| Moderna | Between 1st and 2nd dose | 0.82[6] | Beta (109.1, 25.1) |
| | ≤17 weeks after 2nd dose | 0.94[2] | Beta (125.6, 8.4) |
| | >17 weeks after 2nd dose | 0.92[5] | Beta (125.3, 10.8) |
| Janssen | After 1st dose | 0.66[4] | Beta (55.7, 29.2) |

We used publicly available COVID-19 vaccination data[10] to estimate the weekly number of newly vaccinated individuals in each of the six scenarios and age groups (12-17 years, 18-49 years, 50-64 years, ≥65 years). These age groups were based on vaccine prioritization age groupings. Date of receipt of first and second doses of the Pfizer/BioNTech and Moderna vaccines was not available in our data. We therefore calculated the mean fraction of individuals that received Pfizer/BioNTech or Moderna vaccines in each age group to estimate weekly Pfizer/BioNTech and Moderna vaccinations. We additionally used published literature to estimate the proportion of individuals who received only a single dose of the Pfizer/BioNTech or Moderna vaccines[7].

We combined our estimates of the number of individuals newly vaccinated each week by manufacturer and number of doses received and corresponding vaccine efficacy over time to estimate the fraction of the population with immunity due to vaccination. Since we assumed that natural infection provided perfect immunity, we first calculated the number of newly vaccinated individuals not previously infected with SARS-CoV-2 each week before estimating the number of protected individuals over time due to vaccination. We assumed previously infected and uninfected individuals were equally likely to receive any COVID-19 vaccine, following the observed weekly distribution of vaccines by manufacturer. The formula for calculating the number of new vaccinations in previously uninfected individuals in an age group $a$ at week $t$ is as follows:

$$v_{a,t} = V_{a,t}(1 - \frac{I_{a,t}}{N_{a,t}})$$

$v_{a,t} = \ new\ vaccinations\ in\ susceptible\ individuals$
$V_{a,t} = total\ new\ vaccinations$
$I_{a,t} = number\ of\ unvaccinated\ and\ previously\ infected\ persons$
$N_{a,t} = total\ unvaccinated\ persons$

*Monte Carlo simulation*

We used Monte Carlo simulation to capture uncertainty for the analysis in the alternative model, with a focus on vaccine effectiveness and estimates of asymptomatic infection. We ran 1000 simulations using randomly sampled values of parameters from fitted parameter distributions (Table A1 and A2). We reported the mean and 95% uncertainty intervals (95% UI) of study outcomes.

To generate random samples of our parameters for each simulation, we independently sampled from the distributions of sub-clinical fractions in three age groups: <19 years, 19-59 years, and ≥60 years. We sampled independently from the standard uniform distribution for three vaccines (Pfizer/BioNTech, Moderna, and Janssen) and used inversion sampling to generate samples of vaccine effectiveness to account for changes in effectiveness over time.

Sensitivity analyses

*Age-specific vaccine eligibility*

In both modeling approaches, we performed a sensitivity analysis to account for age-specific differences in COVID-19 vaccine eligibility over time among the four vaccine-eligible age groups (12-17 years, 18-49 years, 50-64 years, ≥65 years).

COVID-19 vaccines became available for the general population ≥16 years and 12-15 years in mid-April 2021 and mid-May 2021 respectively[11]. We assumed vaccination in the population 12-17 years began on April 11, 2021.

Vaccination in adults 18-49 years and 50-64 years began before vaccines were widely available in those populations due to occupational risk. Healthcare and other frontline workers became eligible for COVID-19 vaccines in Phase 1A of vaccination and essential workers became eligible for vaccines in Phase 1B of vaccination[11,12]. For this analysis, we assumed widespread vaccination in the populations 18-49 years and 50-64 years began with Phase 1B of vaccination and used February 14, 2021 to mark vaccine eligibility[13–16].

We used January 10, 2021 as the start of vaccination among adults ≥65 years, since they were eligible for COVID-19 vaccines beginning mid-January 2021[17].

We performed the main analysis in each age group after each age group became eligible for vaccines. Dates of vaccine-eligibility by age group used in this analysis are shown in Table A3.

**Table A3: Date of vaccine-eligibility by age group**

| Age group (years) | Date beginning vaccine-eligibility |
|---|---|
| 12-17 | April 11, 2021 |
| 18-49 | February 14, 2021 |
| 50-64 | February 14, 2021 |
| ≥65 | January 10, 2021 |

*Primary model*

We evaluated the effects of varying the definition of onset of widespread vaccination from November 28, 2020 to January 2, 2021 in weekly timesteps.

We additionally examined the impact of using different age groups (12-17 years, <18 years, <30 years) for the unvaccinated population. Incorporating some vaccine-eligible populations in the unvaccinated population reduced the estimated impact of vaccination.

*Alternative model*
Several variants of SARS-CoV-2 have been identified. Vaccines remained highly effective against infection among most SARS-CoV-2 variants[18,19], but there is evidence of decreased effectiveness of COVID-19 vaccines against symptomatic infection from the delta variant of SARS-CoV-2[18,20,21]. We evaluated study outcomes when considering reduced effectiveness of vaccines against the delta variant after June 2021. The distributions of vaccine effectiveness against clinical disease due to the delta variant are below in Table A4. We did not perform this sensitivity analysis in the primary model since person-level vaccination was not explicitly included.

**Table A4: Mean and fitted parameters for the distribution of possible reduced vaccine effectiveness against the SARS-CoV-2 delta variant**

| Vaccine | Vaccine period | Vaccine effectiveness | |
|---|---|---|---|
| | | Mean | Beta distribution (α, β) |
| Pfizer/BioNTech | Between 1st and 2nd dose | 0.52[3,20] | Beta (12, 11.9) |
| | ≤17 weeks after 2nd dose | 0.95[3,20] | Beta (128.5, 7.2) |
| | >17 weeks after 2nd dose | 0.53[20] | Beta (28.9, 26.3) |
| Moderna | Between 1st and 2nd dose | 0.77[18] | Beta (30.2, 9.5) |
| | ≤17 weeks after 2nd dose | 0.94[2,18] | Beta (125.6, 8.4) |
| | >17 weeks after 2nd dose | 0.80[18] | Beta (71.9, 18.7) |
| Janssen | After 1st dose | 0.60[21] | Beta (10, 7.8) |

14. County A. Alameda County Gets a Mega Vaccination Site through State-Federal Partnership.

### Supplemental tables and figures

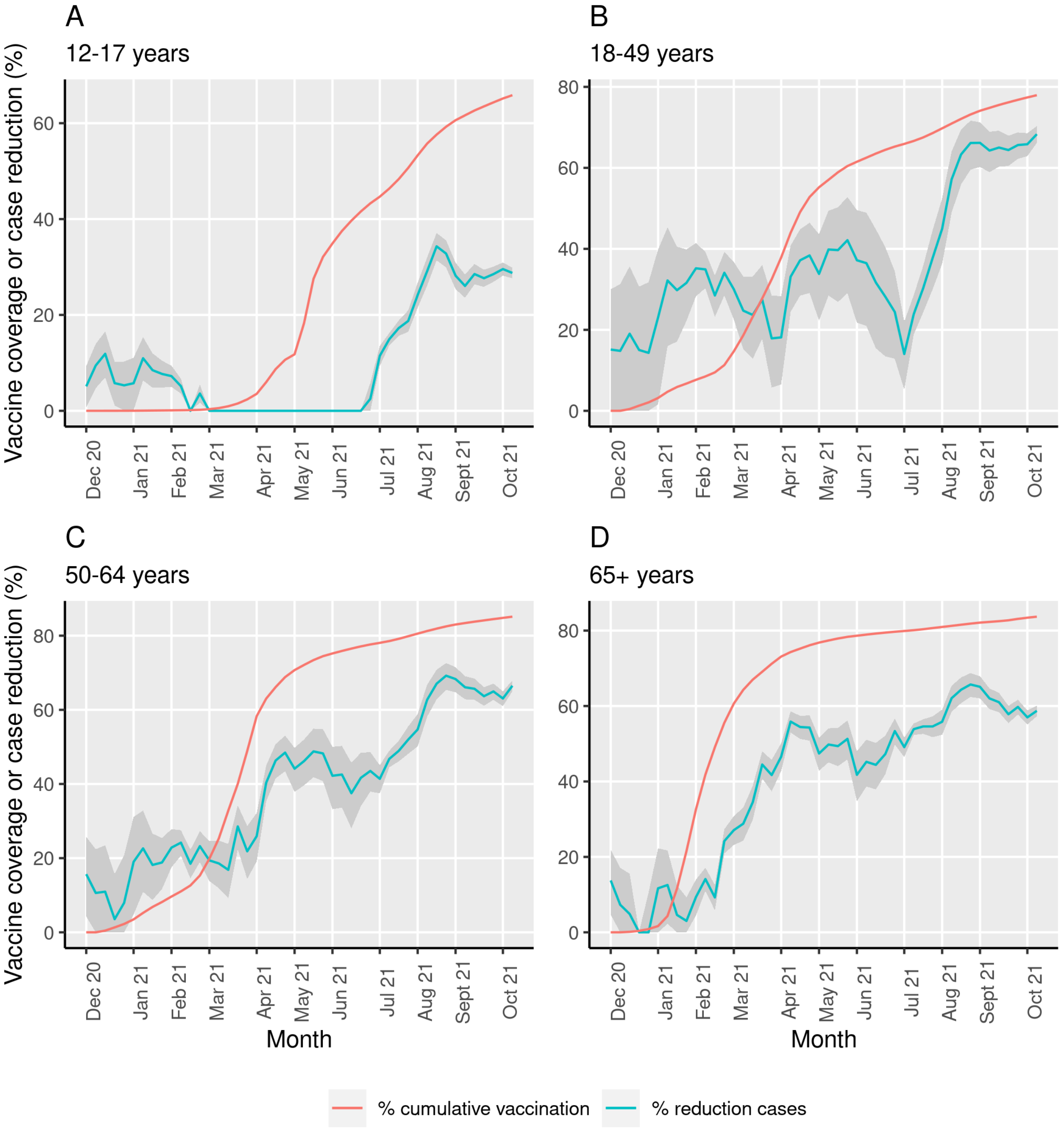

**eFigure 1: Primary model relationship between weekly relative reduction in COVID-19 cases and cumulative vaccination over time**. We estimated the weekly relative reduction in cases due to vaccination in each vaccine-eligible age group (12-17 years in panel A, 18-49 years in panel B, 50-64 years in panel C, ≥65 years in panel D) over the vaccine era (November 29, 2020 - October 16, 2021). We plotted relative reduction in cases (blue) as well as prediction intervals (grey). Cumulative vaccine coverage in each vaccine-eligible age group is shown (red). We identify a relationship between COVID-19 vaccination and relative reduction in COVID-19 cases most clearly in the populations 50-64 years and ≥65 years.

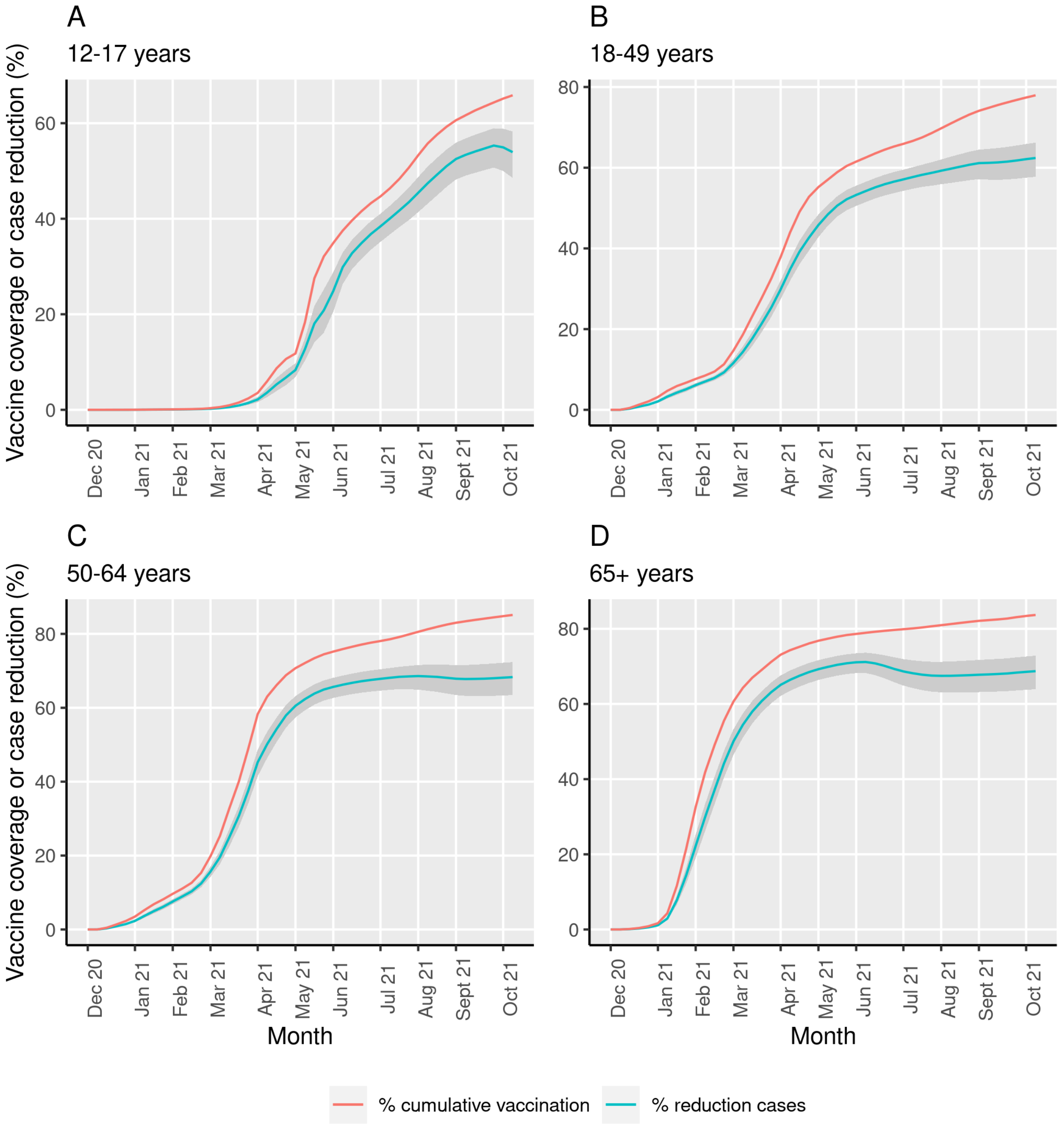

**eFigure 2: Alternative model relationship between weekly relative reduction in COVID-19 cases and cumulative vaccination over time.** We estimated the weekly relative reduction in cases due to vaccination in each vaccine-eligible age group (12-17 years in panel A, 18-49 years in panel B, 50-64 years in panel C, ≥65 years in panel D) over the vaccine era (November 29, 2020 - October 16, 2021). We plotted relative reduction in cases (blue) as well as uncertainty intervals (grey). Cumulative vaccine coverage in each vaccine-eligible age group is shown (red). We identify a proportional relationship between COVID-19 vaccination and relative reduction in COVID-19 cases.

### Sensitivity analyses

#### eTable 1: Sensitivity analysis on age-specific vaccine eligibility on the number of COVID-19 cases averted due to vaccination in California by model and age group

| | | Since Phase 1A | | Since age-based eligibility[a] | |
|---|---|---|---|---|---|
| | Age group (years) | Averted COVID-19 cases (95% PI or UI) | Relative reduction in % (95% PI or UI) | Averted COVID-19 cases (95% PI or UI) | Relative reduction in % (95% PI or UI) |
| **Primary model** | Total (≥12) | 1,523,500 (976,800, 2,230,800) | 34 (25, 43) | 907,280 (804,840, 1,017,760) | 46 (43, 49) |
| | 12-17 | 40,930 (30,270, 52,090) | 15 (11, 18) | 24,340 (22,900, 25,850) | 22 (21, 23) |
| | 18-49 | 1,036,700 (615,100, 1,588,300) | 36 (25, 46) | 554,420 (479,460, 635,510) | 47 (43, 50) |
| | 50-64 | 306,300 (221,500, 415,200) | 34 (27, 41) | 206,160 (189,880, 223,310) | 54 (52, 56) |
| | ≥65 | 139,500 (109,900, 180,200) | 30 (25, 36) | 122,340 (112,600, 133,100) | 43 (41, 45) |
| **Alternative model** | Total (≥12) | 1,402,100 (1,192,100, 1,615,600) | 32 (29, 35) | 1,052,100 (911,300, 1,184,100) | 50 (47, 53) |
| | 12-17 | 78,760 (66,140, 90,610) | 25 (22, 28) | 70,180 (59,590, 79,670) | 45 (41, 48) |
| | 18-49 | 810,700 (697,990, 922,240) | 31 (28, 34) | 592,820 (520,640, 658,400) | 49 (45, 51) |
| | 50-64 | 321,280 (269,390, 375,390) | 35 (31, 39) | 209,990 (182,690, 235,800) | 55 (51, 57) |
| | ≥65 | 191,390 (158,570, 227,340) | 37 (33, 41) | 179,170 (148,330, 210,220) | 53 (48, 57) |

[a]Dates of vaccine eligibility varied by age group. We assumed vaccine eligibility began for 12-17 years on April 11, 2021, 18-49 years on February 14, 2021, 50-64 years on February 14, 2021, and ≥65 years on January 10, 2021.

#### eTable 2: Sensitivity analysis of the primary model measuring averted COVID-19 cases in the vaccine-eligible population when varying the start of the vaccine era

| Start of vaccination | Averted COVID-19 cases (95% PI) | Relative reduction in % (95% PI) |
|---|---|---|
| November 29, 2020 | 1,523,500 (976,800, 2,230,800) | 34 (25, 43) |
| December 6, 2020 | 1,194,900 (925,110, 1,574,770) | 30 (25, 36) |
| December 13, 2020 | 1,151,740 (950,400, 1,408,130) | 31 (27, 36) |
| December 20, 2020 | 1,113,950 (957,140, 1,303,570) | 33 (29, 36) |
| December 27, 2020 | 1,122,320 (974,760, 1,290,440) | 35 (32, 38) |
| January 3, 2021 | 1,129,500 (994,120, 1,272,920) | 38 (36, 41) |

**eTable 3: Sensitivity analysis of the primary model measuring averted COVID-19 cases in vaccine-eligible population using different unvaccinated age groups**

| Unvaccinated population (years) | Vaccine-eligible population (years) | Averted COVID-19 cases (95% PI) | Relative reduction in % (95% PI) |
|---|---|---|---|
| <18 | ≥18 | 1,213,660 (775,470, 1,816,300) | 31 (22, 40) |
| 12-17 | ≥18 | 936,650 (594,770, 1,457,450) | 25 (18, 35) |
| <30 | ≥30 | 231,000 (177,600, 310,450) | 10 (8, 14) |

Note: We evaluated defining the unvaccinated population up to <30 years old, although this population had relatively high vaccination coverage limiting using this age definition for the analysis.

**eTable 4: Sensitivity analysis of the alternative modeling adjusting for possible reduced vaccine effectiveness against the SARS-CoV-2 delta variant**

| Age group (years) | Averted COVID-19 cases (95% UI) | Relative reduction in % (95% UI) |
|---|---|---|
| Total (≥12) | 1,106,300 (913,300, 1,308,650) | 27 (23, 30) |
| 12-17 | 75,190 (62,680, 86,880) | 24 (21, 27) |
| 18-49 | 657,090 (548,600, 768,420) | 26 (23, 30) |
| 50-64 | 241,790 (196,700, 291,550) | 29 (25, 33) |
| ≥65 | 132,190 (105,340, 161,810) | 29 (24, 33) |

We incorporated estimates of vaccine effectiveness against the delta variant after June 2021 (see Table A4) in the main analysis.
